# Estimating the correlation between rates of change in longitudinal studies in presence of dropout

**DOI:** 10.64898/2026.10.16.26365737

**Authors:** Ruben P. Dörfel, Brice Ozenne, the Alzheimer’s Disease Neuroimaging Initiative (ADNI)

## Abstract

Estimating the correlation between subject-specific rates of change is a common objective in longitudinal studies. A natural approach is to fit ordinary least squares regressions per subject and correlate the resulting slope estimates directly. Under complete data, this slopes-first estimator is algebraically equivalent to a contrast-based plug-in estimator derived from the joint variance-covariance matrix of the two signals across time. Under independent dropout, however, the slopes-first approach implicitly applies heterogeneous weights across missing-data patterns, introducing bias whenever the cross-covariance between the two signals is not constant over time. We formalise this result and propose an unbiased alternative that estimates the joint covariance matrix before applying the contrast. We further extend our results to include covariates as well. Simulations confirm that the slopes-first estimator is biased under non-constant cross-covariance while the proposed alternative remains unbiased. Further, we show in a real-world example that the two estimators can lead to different scientific conclusions and recommend that researchers working with longitudinal data inspect the missing data pattern to make an informed decision about the estimator of choice.

## 1 Introduction

In longitudinal neuroimaging studies, researchers often seek to understand whether changes in one brain measure are associated with changes in another. For instance, the rate of grey matter atrophy may correlate with the rate of cognitive decline, or accelerated brain aging, as measured by brain-predicted age deviation, may track with structural changes over time [1]. Other examples could be potential association between change in tau and amyloid-*β* over time [2] or whether a change in dopamine D2/D3 receptor binding was associated with a change in depression symptoms in response to therapy [3]. Answering such questions requires estimating the correlation between subject-specific rates of change across two longitudinal measures. Accurately quantifying these relationships is essential for identifying disease mechanisms, validating biomarkers, and selecting endpoints for clinical trials. One option would be to estimate the absolute change, i.e., the difference in first/last. However, those measures are typically measured with noise. If intermediate values are available, estimating the rate of change, or slope, could help to get a more accurate estimate of the change. Studies with planned and therefore common measurement times across subjects are recommended for this purpose, as otherwise assumptions about the association between the brain measures and visit processes needs to be made [4]. This design is widely used in studies with multiple timepoints at fixed intervals. Common examples of such studies are ADNI [5], OASIS [6], PPMI [7], or ABCD [8].

For illustrative purposes, we will consider the analysis in [1] aiming to relate a change in brain predicted age (*X*) with a change in grey matter volume (*Y*). Each of the *N* = 666 cognitively normal participants was followed for up to four years with study visits at each of the *T* = 5 timepoints (including baseline at *t* = 0). Let *x*_*i,s*_ and *y*_*i,s*_ denote the two brain signals, where *i* ∈ {1, …, *T*} indexes timepoints and *s* ∈ {1, …, *N* }indexes subjects. For each subject, the rate of change in brain age and grey matter volume were estimated by the slopes *β*_*x,s*_ and *β*_*y,s*_ from linear regressions of *x*_*i,s*_ and *y*_*i,s*_ on time *t*. The scientific objective was to determine whether the change in brain age and change in grey matter volume were correlated across subjects. More formally, we wish to estimate the Pearson correlation coefficient

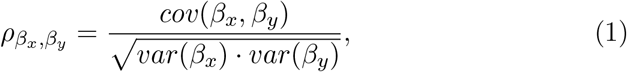

where *β*_*x*_ and *β*_*y*_ are random variables whose realizations are the subject-specific slopes *β*_*x,s*_ and *β*_*y,s*_ obtained using ordinary least squares. A related problem involves correlating a baseline value *x*_1_ with a slope *β*_*y*_ to examine whether an initial status is associated with the rate of change.

A straightforward approach is to perform ordinary least squares separately for each subject to obtain subject-specific estimates of the slopes, denoted 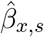 and 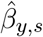, and then estimate their correlation:

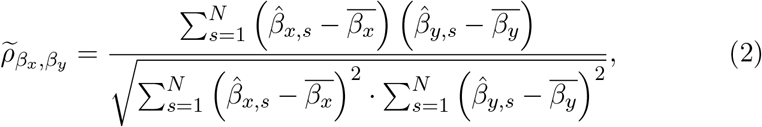

where we use the bar notation to denote the empirical averages: 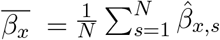 and 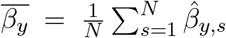 . With complete data, we will show that this estimator can be re-written as a weighted average of the covariance between the two signals *x* and *y* at each of the *T* ^2^ pairs of timepoints. However, longitudinal studies invariably suffer from missing data due to dropout, missed visits, or study completion before all timepoints are collected. One such example with data taken from [1] is presented in Table 1, where 290 of 666 participants (44%) entering the study remained until four years from baseline, or in other words, 56% were missing at the end of the study.

**Table 1.** Cognitively normal subjects scanned at each timepoint. Missing [%] is the percentage of baseline subjects not scanned at that timepoint.

| Years from Baseline | 0 | 1 | 2 | 3 | 4 |
| --- | --- | --- | --- | --- | --- |
| Subjects Scanned | 666 | 453 | 518 | 197 | 290 |
| Missing [%] | 0% | 31.98% | 22.22% | 70.42% | 56.45% |

In presence of missing data, the slopes-first strategy may seem suboptimal as subjects with longer follow-up should provide more precise estimated slopes, yet the standard correlation calculation fails to account for this heterogeneity in estimation precision. We demonstrate that the problem is more acute: the slopes-first strategy can produce biased estimates of 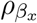, *β*_*y*_ . Indeed the weights used when pooling the time-specific covariance between the two signals to estimate 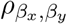, depend on the number of available subjects at those times. Missing data, and especially dropout, lead to a different weighting scheme if the association between the two signals varies over time. We introduce an alternative method for computing the correlation between two measures that is unbiased when data are either missing completely at random (MCAR) or missing at random (MAR), i.e. when dropout depends only on previously observed values. We then use a simulation study to confirm these claims and illustrate the use of the two estimators on a real-world case study. Finally, we show how to adapt the methodology to estimate partial correlation, e.g., correlation between the slopes adjusted for baseline values.

## 2 Methods

We represent the two signals between time 0 and time *τ* by two stochastic processes (*X*_*i*_)_*i*∈[0,*τ*]_ and (*Y*_*i*_)_*i*∈[0,*τ*]_, assumed to have finite mean, finite variance, and positive definite finite covariance. The two processes are monitored in discrete time over a set of timepoints *T* denoted of size *T* . In the application *τ* = 4 years and measurement times are recorded in years so *T* can be at most 5.

The *T* × *T* matrices **Σ**_*XX*_ and **Σ**_*Y Y*_ will denote the variance-covariance matrices of, respectively, (*X*_*i*_)_*i*∈*T*_ and (*Y*_*i*_)_*i*∈T_ . For *i, j* ∈ *T*, their diagonal elements will be denoted 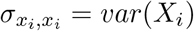and 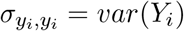, and their off-diagonal elements 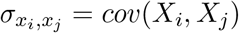 and 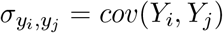, respectively. The *T* × *T* matrix **Σ**_*XY*_ will denote the cross-covariance matrix between (*X*_*i*_)_*i*∈i_ and (*Y*_*i*_)_*i*∈*T*_, with entries 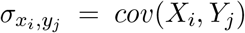 for *i, j* ∈ *T*, and 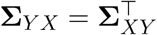 .

The joint covariance matrix between the two signals and over the measurement timepoints will be denoted 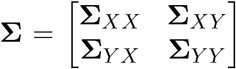 while the covariance matrix between the slopes will be denoted 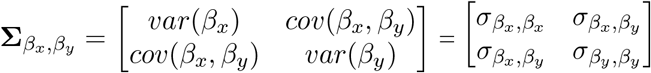.

### 2.1 Complete data case

We first consider the case where a measurement is available for each subject at each of the planned measurement times: *T*= (0, 1, 2, 3, 4) and *T* = 5. We now explicit the relationship between the estimated parameter of interest, 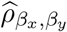, and some of the estimated characteristics of the signals, 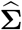.

We start by expressing 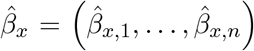 and 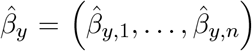 as a function of time and the observed variables. Following standard results, e.g., chapter 1 of [9], for a given subject *s* ∈ {1, …, *N* }, the ordinary least squares estimate of the slope in the linear regression *y*_*i,s*_ = *α*_*y,s*_ + *β*_*y,s*_*t*_*i*_ + *ε*_*i,s*_:

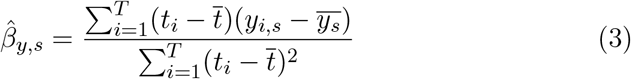

where we used the bar notation to express the empirical average over time. The formula for 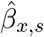 follows by symmetry between *x* and *y*. Since the times *t*_*i*_ are the same for all subjects, we show in appendix A.1 that the estimated slope can be re-written as a weighted average of the measured signal over time:

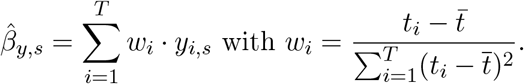

The weight *w*_*i*_ only depends on the timepoints and their number *T* but not on the signal value. So if we let **w** ∈ ℝ^*T* ×1^ be the vector containing all weights *w*_*i*_ and **Y, X** ∈ ℝ^**T**×**N**^ the matrices containing all values *y*_*i,s*_ and *x*_*i,s*_ respectively, we therefore express 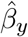and 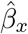 both as 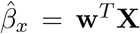 and 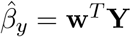 .

Noticing that equation (1) only involves elements of the variance-covariance matrix 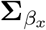, *β*_*y*_, it only remains to explicit 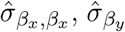, β_*y*_ and 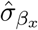, β_*y*_ . We show in appendix A.3 that the variance of *β*_*y,s*_ can be expressed as:

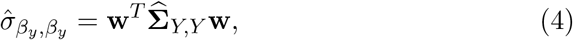

with 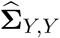 being the estimated covariance matrix across time points. The variance of *β*_*x,s*_ follows by symmetry between *x* and *y*. Finally, as shown in appendix A.4, the covariance between 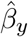 and 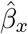 can be expressed as

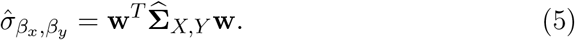

Putting things together, we show in appendix A.5 that the plug-in estimator for the correlation based on formula (1) can be expressed as:

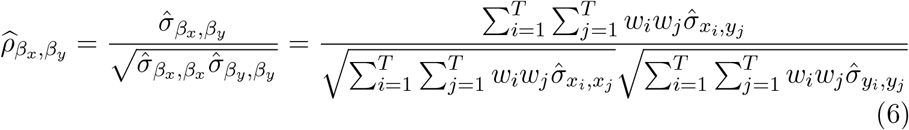

which is equal, in the special case of complete data, to 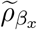, *β*_*y*_the slopes-first estimator strategy (appendix A.6). When correlating a baseline value *x*_1_ with a slope *β*_*y*_, (6) would simplify to

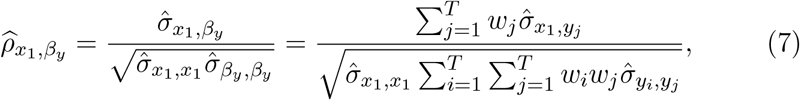

as shown in the appendix (A.18). If the covariance between the baseline measurement and the other signal is constant over time 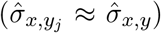, the estimated correlation will be 0 as the weights sum to 0.

To conclude, we show two different strategies to estimate the correlation 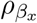, *β*_*y*_ :

- *Strategy 1* : estimate *β*_*x,s*_ and *β*_*y,s*_ separately for each subject via ordinary least squares, and then compute their correlation via equation (2).
- *Strategy 2* : estimate the covariance matrix **Σ** between the two signals across time. Compute the weights 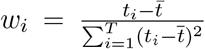 .Estimate the correlation by evaluating weighted averages of the covariance according to equation (6).

Strategy 1 avoids to estimate the joint variance-covariance matrix but requires to estimate the subject specific slopes. From a computational point of view, Strategy 2 is preferable when (*N* ≫ *T*) while Strategy 1 is preferable when (*T* ≫ *N*).

### 2.2 Incomplete data case

As mentioned in the introduction, not all subjects have an observation at each of the possible measurement timepoints, possibly because they dropped out of the study, or missed a visit. In appendix A.7, we show that, in the case of independent dropout, estimating each slope separately *(Strategy 1)* will lead to biased estimates of the overall correlation. More specifically, the correlation estimator can be understood as a ratio of weighted average of covariances:

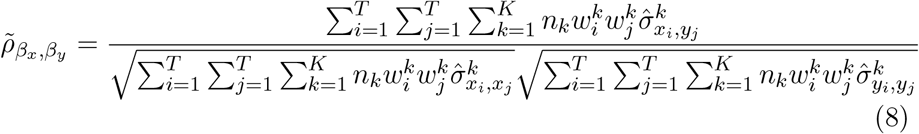

Denoting by *K* the number of missing data patterns, by 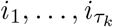 the index of the timepoints at which measurements were taken in the missing data pattern *k*, and by 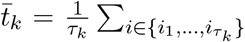 the average measurement time in this missing pattern, the weights can be expressed as:

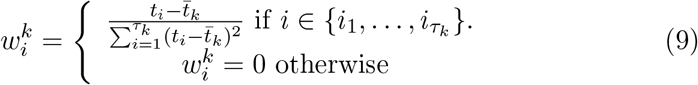

*Strategy 1* (slopes-first) weights each pair of timepoints depending on the available data via the weights 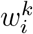 and 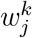 at these timepoints, while *Strategy 2* uses pre-defined weights that only depend one the measurement times: *w*_*i*_ and *w*_*j*_ in equation (6).

In presence of missing observations, there is not a unique way to estimate the joint variance-covariance matrix for Strategy 2. We consider two possible estimators:

- *Strategy 2a*: estimate 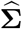 using only subjects with complete data across all timepoints, constructing a complete-case analysis. This approach is similar to using the function `cov()` with pairwise or complete case options in R or Python.
- *Strategy 2b*: use a linear mixed model to estimate 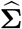 . First stack **X** and **Y** into a single vector denoted **Z** = {*X*_1_, …, *X*_*T*_, *Y*_1_, …, *Y*_*T*_ }. A linear mixed model with non-parametric mean and covariance structure for **Z** fitted with Restricted Maximum Likelihood (REML) will provide a consistent estimator for **Σ**. In statistical software this is typically obtained using a categorical variable indexing the name of variable from which the **Z** value originates as regressor and an unstructured covariance matrix.

REML is unbiased in special cases (section 3.6.1 [10]); otherwise its bias has been shown to decay at a quadratic rate with the sample size, improving over Maximum Likelihood where it decays at a linear rate [11]. The two are known to be asymptotically equivalent (section 3.6.3 of [10] for a proof with a simpler covariance pattern) meaning that REML is consistent when missingness in **Z** is either completely random (MCAR) or only depends on elements in **Z** (MAR) and elements in **Z** are linearly related (section 14.2 of [12]).

While straightforward to implement, *Strategy 2a* estimates each covariance element separately and therefore does not use the information in incomplete trajectories efficiently. When using the pairwise option, the resulting matrix is not guaranteed to be positive semi-definite, and the estimates are not guaranteed to be consistent when dropout depends on previously observed values (MAR). *Strategy 2b* incorporates information from all subjects, including those with incomplete follow-up, in a single likelihood. Under the assumption of independent dropout, Strategy 2b should provide more accurate estimates of 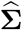 and consequently of 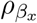, *β*_*y*_, as it makes fuller use of the available information, and it remains consistent under MAR.

### 2.3 Estimating partial correlation

Strategy 1 and 2 can be modified to control for covariates *Z*, assumed *Q* dimensional with *Q < T* . This is particularly relevant when baseline values may confound the relationship between changes over time, and when individuals starting at different baseline levels may have systematically different capacities for change due to ceiling or floor effects.

- *Strategy 1* : estimate the covariate adjusted slopes, denoted *β*_*x,s*|*z*_ and *β*_*y,s*|*z*_, using a linear regression regression *X* and *Y* against time and the covariates *Z*. Then compute the partial correlation (between slopes) by plugging *β*_*x,s*|*z*_ and *β*_*y,s*|*z*_ in place of *β*_*x,s*_ and *β*_*y,s*_ in formula (2). This estimator will be denoted 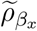, *β*_*y*_|*z*.
- *Strategy 2* : estimate the covariance matrix 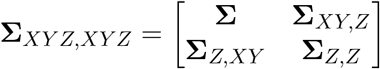 between the two signals across time and covariates. Compute the weights 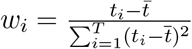 and the conditional covariance matrix 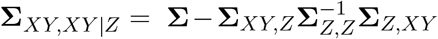 . Deduce the correlation by evaluating weighted averages of the conditional covariance by plugging them in place of (marginal) variances 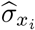,*x*_*j*_,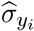, *y*_*j*_ and 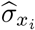, *y*_*j*_ in formula (6).

Equivalence between two strategies in the complete data case is shown in appendix A.6. The derivation of Strategy 2 is provided in A.8 in the special case where we adjust for baseline signal value: *Z* = (*X*_1_, *Y*_1_). The derivations involve the 4 × 4 variance-covariance matrix of the two changes with their baseline variables and deriving the conditional variance-covariance matrix of the changes given baseline using the standard formula for conditional distributions in multivariate normal theory. In this special case the double sum in formula (6) could start at 2 instead of 1 as the conditional variances are 0 for *i* = 1 or *j* = 1.

## 3 Experiments

In what follows, we will use simulation studies to quantify the bias of Strategy 1 in the case of independent dropout similar to our motivating example, as well as in the case of dropout depending on previously observed values. We will further show that this holds when adjusting for baseline values, and compare different estimation strategies for the joint variance-covariance matrix. Further, we provide a real-world example showing that the different strategies can indeed lead to different conclusions. All code to reproduce the simulations is publicly available at https://github.com/RDoerfel/bap1b-corr-public.

### 3.1 Simulations

To verify that estimating the correlation via the joint variance-covariance matrix (*Strategy 2*) leads to unbiased estimates in the presence of missing data, we simulated longitudinal data with dropout and investigated three different cross-correlation patterns (see Figure 1). For a given joint covariance matrix **Σ** and mean ***µ*** = (***µ***_*X*_, ***µ***_*Y*_), we simulated 1000 datasets of 1000 subjects each from a multivariate normal distribution

**Figure 1.**
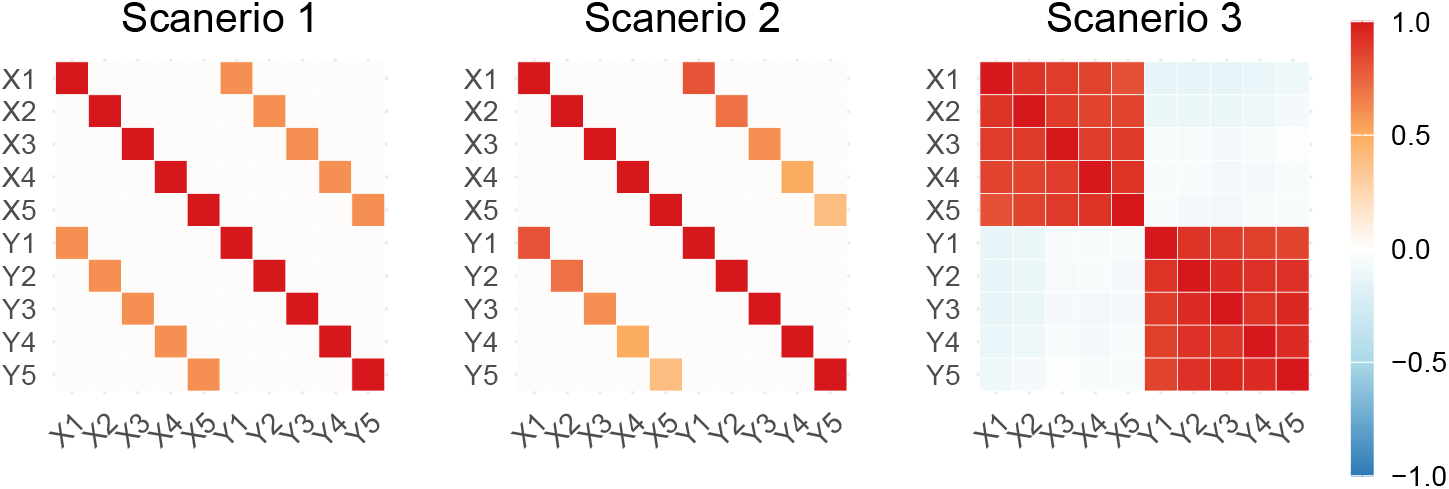
Joint variance-covariance structures used for the simulations: **Scenario 1)** Constant diagonal **Σ**_**X**,**Y**_; **Scenario 2)** Non-constant diagonal **Σ**_**X**,**Y**_; **Scenario 3)** realistic **Σ** estimated from real data.

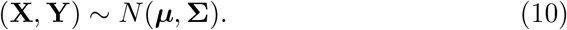

Dropout was simulated as monotone attrition similar to the observed retention counts in Table 1. Even though we observed a mix of missingness and monotone attrition we, for simplicity, only consider monotone attrition in our simulations. For each simulated subject, a last observed timepoint was drawn from {0, 1, 2, 3, 4 }with probabilities (0.22, 0.10, 0.24, 0.14, 0.30), corresponding to cumulative missingness of 22%, 32%, 56% and 70% at years 1–4. This is a stylised monotone approximation of Table 1, in which the missing percentages were sorted to be non-decreasing. Here, dropout is independent of the data, i.e. missing completely at random (MCAR). To also assess dropout that depends on previously observed values, i.e. missing at random (MAR), we repeated each scenario with the same expected retention, but let the probability of dropping out after visit *t* depend on the subject’s observed change from baseline, logit *P* (dropout after *t*) = *a*_*t*_ + *βz*_*t*_, where *z*_*t*_ is the standardised change *Y*_*t*_ −*Y*_0_ and *β* = −2, so that subjects with a larger decline in *Y* are more likely to drop out. The intercepts *a*_*t*_ were calibrated in each dataset so that the expected retention matches the MCAR case. We refer to these MAR scenarios as Scenarios 1b–3b. Given the predefined joint covariance matrix **Σ** and equation (6), we computed the theoretical correlation (1) before substituting NAs to the signal values according the simulated dropout and compared it against the estimates from 1000 Monte Carlo replicates. For each simulated dataset, we estimated the correlation between slopes using all three strategies. Strategy 1 computes slopes separately for each subject using available data, then correlates these estimated slopes directly. Strategy 2a estimates the joint covariance matrix 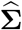 using pairwise complete observations, i.e. each covariance element is estimated from the subjects observed at both of the respective timepoints, then applies the contrast-based approach. Strategy 2b estimates 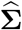 using a linear mixed model that incorporates all available data, including subjects with incomplete follow-up, before applying the same contrast-based computation. This comparison allows us to assess both the bias introduced by the slopes-first approach and the efficiency gains from using all available data via mixed models. Additionally, we estimated the correlation between slopes using all three strategies adjusted for the respective baseline values as described in 2.3, and more specifically the appendix A.8.

We considered three scenarios with distinct covariance structures, visualized in Figure 1. In the first scenario, **Σ**_*X,Y*_ is a diagonal matrix with constant cross-covariance 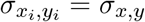 at all timepoints. In the second scenario, **Σ**_*X,Y*_ remains diagonal but with varying diagonal elements 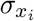,*y*_*i*_ across time. In the third scenario, we estimated both ***µ*** and **Σ** directly from real data to capture realistic covariance structures including off-diagonal elements. For the first two scenarios, **Σ**_*X,X*_ and **Σ**_*Y,Y*_ were set to identity matrices to isolate the effect of different cross-covariance patterns on bias.

We evaluated the simulations following [13]. For *n*_sim_ replicates with estimates 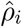 and true value *ρ*, the bias is 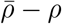, where 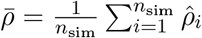 It measures whether a strategy is, on average, centred around the truth. The empirical standard error, 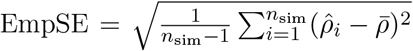, measures the precision of the estimator, i.e. the spread of 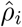 across replicates. The root mean squared error, 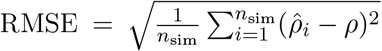, combines both. To quantify the uncertainty due to the finite number of replicates, we report Monte Carlo standard errors (MCSE) for the bias, 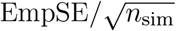, and for the empirical standard error, 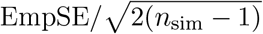 [13]. Replicates in which model fitting failed were excluded, so *n*_sim_ may be slightly less than 1000.

The results are presented in Figure 2 and Table 2. In agreement with the derivations made in appendix A.6, Scenario 1 under MCAR is a special case where all strategies lead to similar results. However, as soon as the cross-term of the joint variance-covariance matrix are not constant, i.e. the correlation between measurements vary across timepoints, Strategies 2a and 2b led to estimates centred around the ground truth whereas Strategy 1 was notably biased (bias of 0.113 and 0.059 in Scenarios 2 and 3). Under MAR dropout, the bias of Strategy 1 increased further (0.165 and 0.147 in Scenarios 2b and 3b) and was also present in Scenario 1b (0.049), whereas Strategy 2b remained unbiased in all scenarios. Strategy 2a was approximately unbiased in Scenarios 1 and 2, but in the realistic Scenario 3 its empirical standard error was about four times that of Strategy 2b (0.197 vs. 0.050 under MCAR and 0.221 vs. 0.056 under MAR), with a small bias under MAR (0.023). The same pattern was observed with baseline adjustment (Fig. B.1).

**Table 2.** Simulation results by scenario, missingness mechanism, and strategy, based on 1,000 Monte Carlo replicates. True *ρ* is the theoretically derived value and Mean 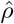 is the mean estimated correlation across replicates. Bias is Mean 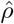minus True *ρ*. Emp. SE is the empirical standard error (the standard deviation of 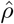 across replicates), and RMSE is the root mean squared error. Monte Carlo standard errors are given in parentheses.

| Scenario | Mechanism | Strategy | True $\rho$ | Mean $\hat{\rho}$ | Bias | Emp. SE | RMSE |
| --- | --- | --- | --- | --- | --- | --- | --- |
| S1 | MCAR | 1 | 0.600 | 0.599 | −0.001 (0.001) | 0.037 (0.001) | 0.037 |
|  |  | 2a | 0.600 | 0.597 | −0.003 (0.001) | 0.035 (0.001) | 0.035 |
|  |  | 2b | 0.600 | 0.597 | −0.003 (0.001) | 0.035 (0.001) | 0.035 |
| S2 | MCAR | 1 | 0.600 | 0.713 | 0.113 (0.001) | 0.027 (0.001) | 0.117 |
|  |  | 2a | 0.600 | 0.596 | −0.004 (0.001) | 0.036 (0.001) | 0.036 |
|  |  | 2b | 0.600 | 0.596 | −0.004 (0.001) | 0.036 (0.001) | 0.036 |
| S3 | MCAR | 1 | −0.170 | −0.111 | 0.059 (0.002) | 0.049 (0.001) | 0.077 |
|  |  | 2a | −0.170 | −0.179 | −0.009 (0.006) | 0.197 (0.004) | 0.197 |
|  |  | 2b | −0.170 | −0.173 | −0.003 (0.002) | 0.050 (0.001) | 0.050 |
| S1b | MAR | 1 | 0.600 | 0.649 | 0.049 (0.001) | 0.034 (0.001) | 0.060 |
|  |  | 2a | 0.600 | 0.599 | −0.001 (0.001) | 0.031 (0.001) | 0.031 |
|  |  | 2b | 0.600 | 0.599 | −0.001 (0.001) | 0.035 (0.001) | 0.035 |
| S2b | MAR | 1 | 0.600 | 0.765 | 0.165 (0.001) | 0.023 (0.001) | 0.166 |
|  |  | 2a | 0.600 | 0.599 | −0.001 (0.001) | 0.033 (0.001) | 0.033 |
|  |  | 2b | 0.600 | 0.599 | −0.001 (0.001) | 0.037 (0.001) | 0.037 |
| S3b | MAR | 1 | −0.170 | −0.022 | 0.147 (0.002) | 0.052 (0.001) | 0.156 |
|  |  | 2a | −0.170 | −0.147 | 0.023 (0.007) | 0.221 (0.005) | 0.222 |
|  |  | 2b | −0.170 | −0.169 | 0.000 (0.002) | 0.056 (0.001) | 0.056 |

**Figure 2.**
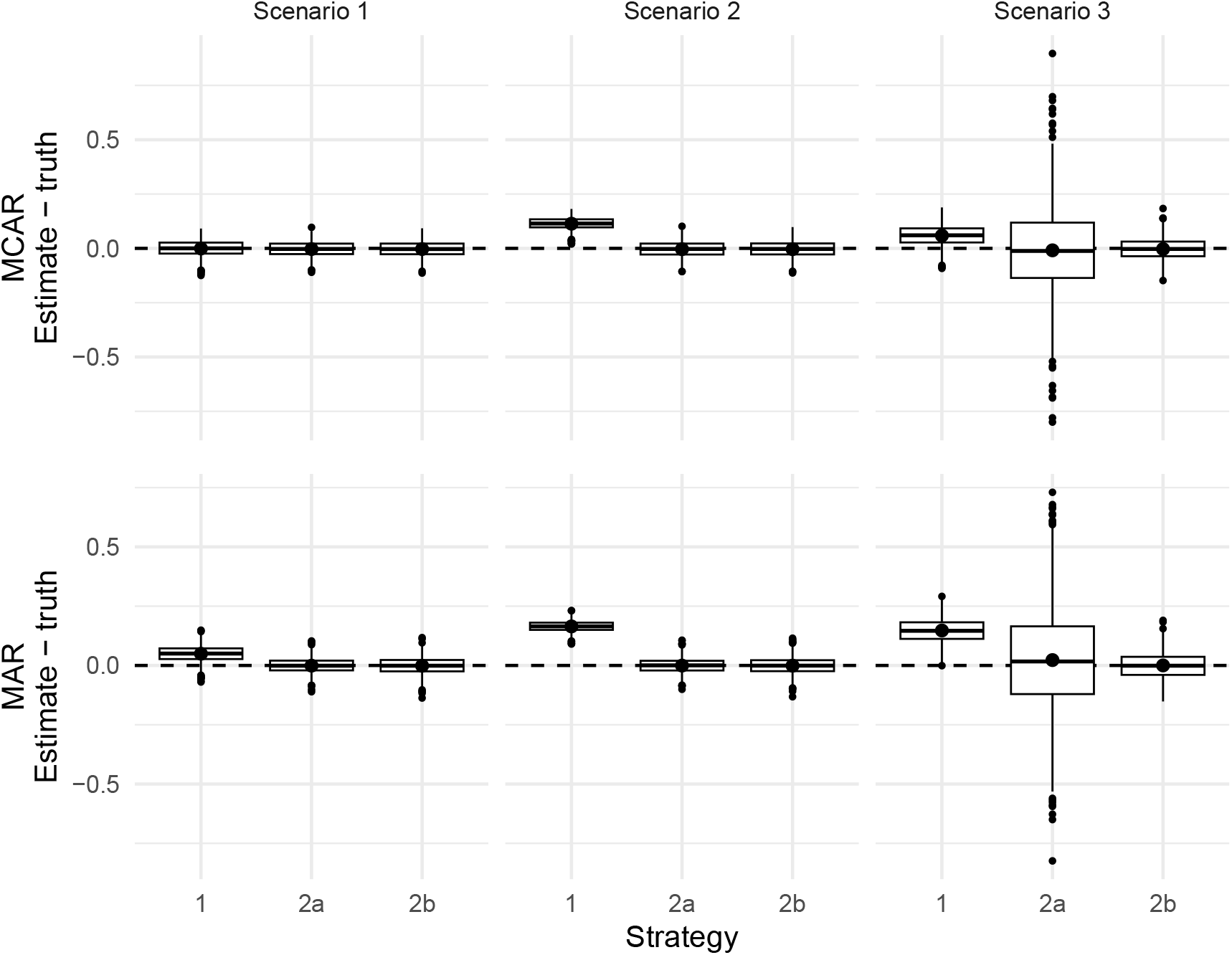
Error in estimated correlations between slopes (estimate-truth) across three covariance structures (Scenarios 1–3), without baseline adjustment. Top row: missing completely at random (MCAR); bottom row: missing at random (MAR) dropout. Strategy 1: slopes-first approach; Strategy 2a: joint covariance approach via pairwise complete case; Strategy 2b: joint covariance, approach via linear mixed model. The dashed line marks zero error (no bias). Boxplots show 1000 Monte Carlo replicates. The points show the mean error. Results with adjustment are shown in Fig. B.1

### 3.2 Case Study: Brain Age and grey Matter Volume

To illustrate the two strategies on real data, we revisit a case study from our previous work [1]. There, we investigated whether changes in brain-predicted age (brain age) are correlated with changes in grey matter volume, i.e., we aimed to estimate *ρ* as defined in eq. (1). The Data used in the preparation of this article were obtained from the Alzheimer’s Disease Neuroimaging Initiative (ADNI) database (`adni.loni.usc.edu`). In short, the ADNI has acquired a longitudinal dataset over multiple years where they acquired, among other things, structural MRI scans for subjects with normal cognition, mild cognitive impairment, and Alzheimer’s disease. Over time, subjects drop out or miss scans. Here we include data up to four years after baseline, meaning we consider five timepoints in total. We used pyment to predict brain age [14], and SPM12 [15] to estimate the cerebral grey matter volume. Finally, we estimated the correlation between the change across the five timepoints according to the three different strategies (1, 2a, 2b). More details about the motivation of the analysis can be found in Dörfel et al. [1].

We report point estimates of 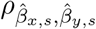 along with their 95% confidence intervals (CIs) for all three strategies in Table 3. The CIs and p-values for *Strategy 1* were estimated by the linear model, those for Strategy 2a by bootstrapping, and those for Strategy 2b by the delta method. We further provide baseline-adjusted correlations using the methods described in Appendix A.8, which control for initial differences in brain age and grey matter volume when estimating the correlation between rates of change. Strategy 1 estimated a negligible correlation, while Strategy 2b estimated a correlation roughly five times larger in magnitude and a substantially lower p-value (0.0623 instead of 0.5777). Strategy 2a had a broad confidence interval, which we attribute to the small number of participants with complete data which are the only one used by this strategy. The difference between the results from Strategy 1 and 2b can be explained by the weights used by the different analyses. They are shown in Figure 4. In brief, Strategy 1 put a greater emphasis on earlier timepoints while Strategy 2 mostly weight the first and last timepoints, similarly to when correlating the change from baseline, and no weight in the ‘middle’ timepoint. Together with the joint variance covariance matrix (see Figure 3), it is possible to apply eq. (6). For example, the numerator in this equation could be computed by multiplying the lower left block of the joint variance-covariance matrix in Figure 3 by the weights visualized in 4b. A more accessible case, is derived in the appendix A.7.

**Table 3.** Correlation between the rates of change in brain-predicted age deviation and grey matter volume over four years for Strategies 1, 2a and 2b, unadjusted and adjusted for baseline values. Point estimates are accompanied by 95% confidence intervals (CIs) and their p-value.

| Strategy | Estimate | 95% CI Lower | 95% CI Upper | p-value |
| --- | --- | --- | --- | --- |
| 1 | -0.022 | -0.099 | 0.0553 | 0.5777 |
| 2a | 0.0376 | -0.3573 | 0.4326 | 0.8518 |
| 2b | -0.1208 | -0.248 | 0.0065 | 0.0623 |
| 1 adjusted | -0.0023 | -0.0735 | 0.0689 | 0.9498 |
| 2a adjusted | 0.0557 | -0.2911 | 0.4025 | 0.7528 |
| 2b adjusted | -0.1206 | -0.2488 | 0.0076 | 0.0645 |

**Figure 3.**
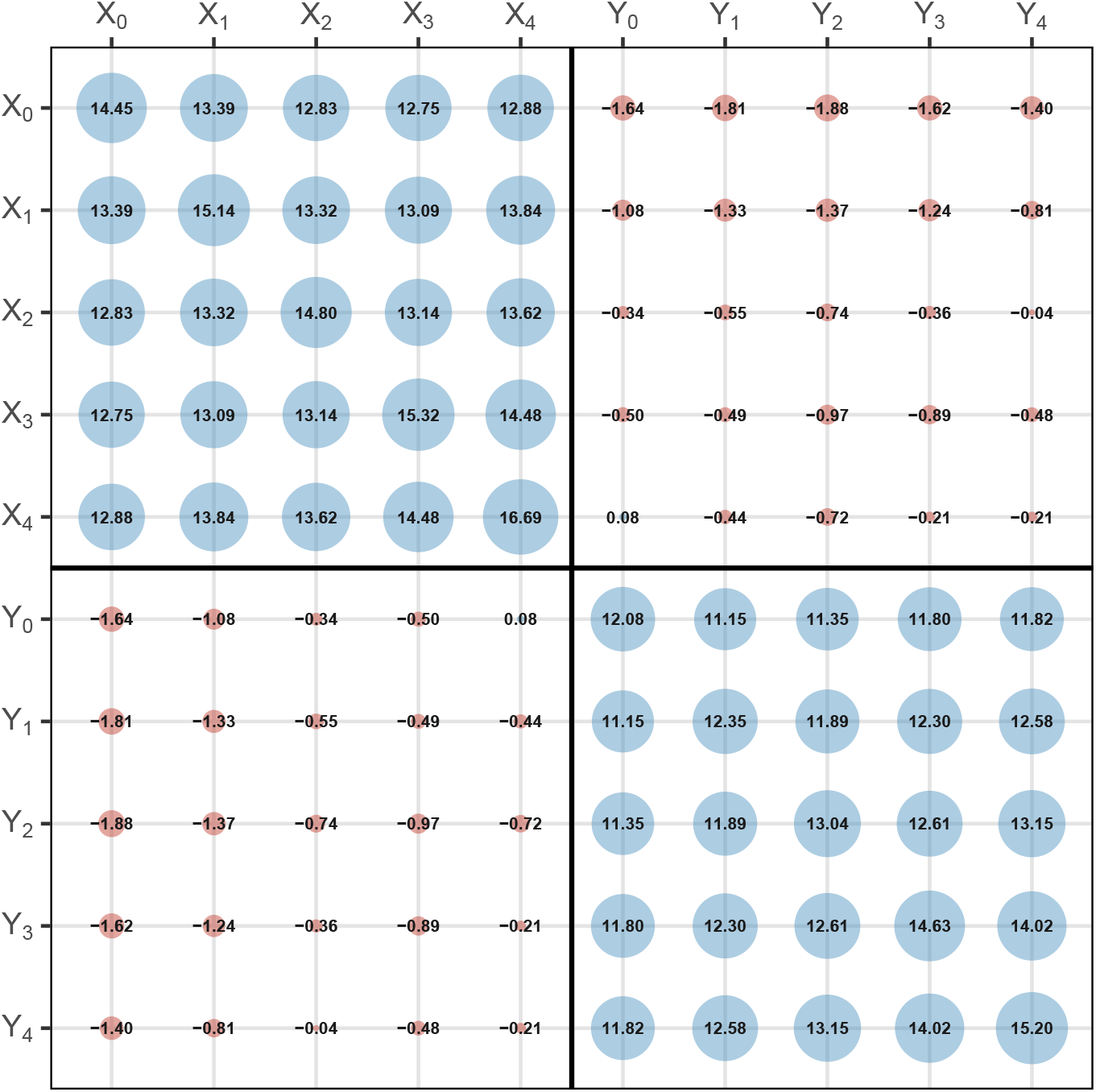
Estimated joint variance-covariance matrix **Σ** estimated from the real data. The size of the circles is proportional to the value, displayed at the center of each dot, in the joint matrix and the colour indicates the sign (red: negative; blue: positive).

**Figure 4.**
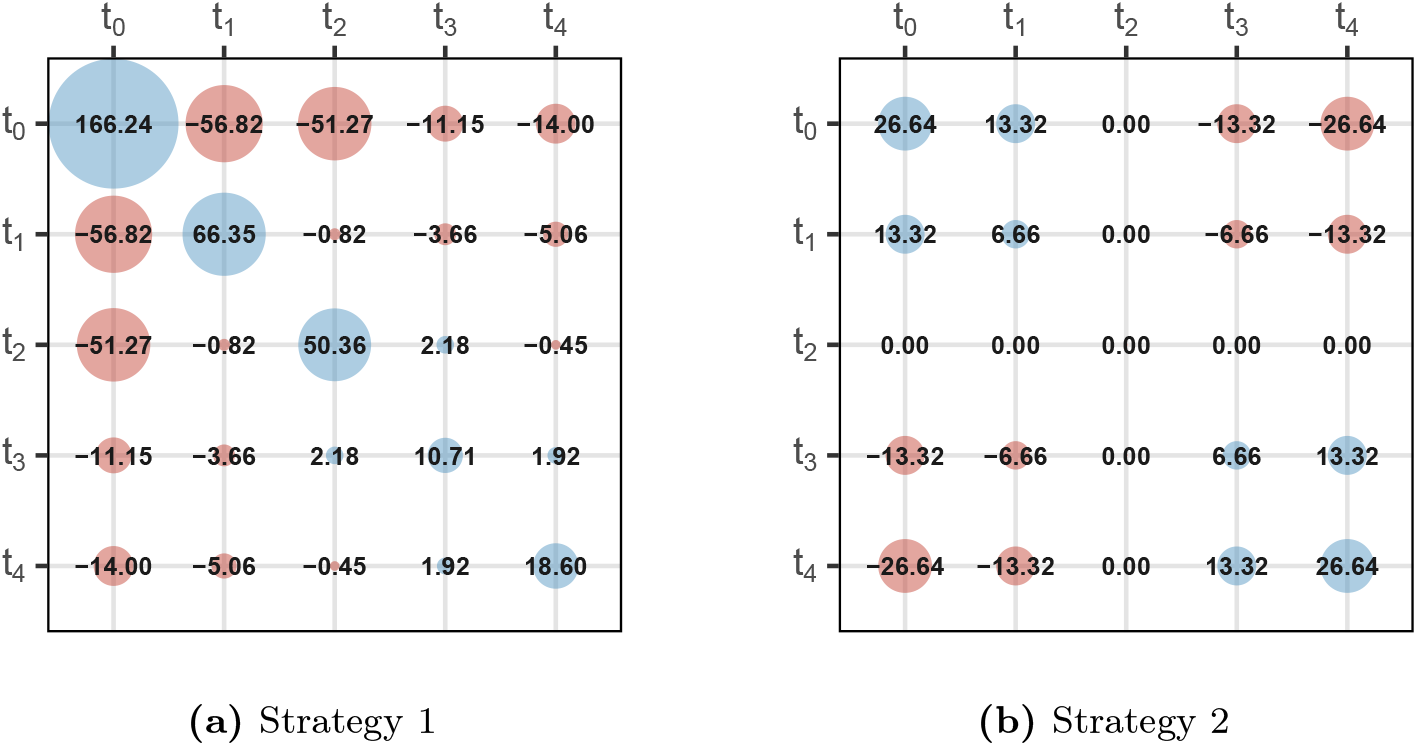
Weighting of each 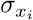,*y*_*j*_ displayed in Figure 3 for the respective time pairs in each of the four blocks. The value at the centre of each circle is 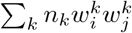 for Strategy 1 (a) and *Nw*_*i*_*w*_*j*_ for Strategy 2 (b). The size of the circles is proportional to the maximum absolute value in both weight matrices, and the colour indicates the sign (red: negative; blue: positive).

## 4 Discussion

Our analysis demonstrates that the slopes-first approach (Strategy 1) for estimating correlations between longitudinal trajectories can produce biased results in the presence of missing data whenever the cross-covariance structure varied across timepoints. Notably, this bias persisted even under the favourable assumption of independent dropout, and was evident in both simplified simulation scenarios and when using realistic covariance structures derived from neuroimaging data.

Under complete data, the two strategies are algebraically equivalent (Appendix A.6). Likewise, when the cross-covariance between the two signals is constant across timepoints, and dropout is independent (MCAR), as in Scenario 1 of our simulations, Strategy 1 remains unbiased, because the heterogeneous per-subject weights introduced by dropout cancel in the ratio. However, this no longer holds when dropout depends on the observed values (Scenario 1b), since the subjects with fewer timepoints, and thus different weights, then also differ systematically in their slopes. In practice, neither the assumption of a time-constant cross-covariance nor that of independent dropout is verifiable a priori. Strategy 2 is therefore preferable whenever incomplete follow-up with potentially dependent dropout is present.

Within Strategy 2, the choice of covariance estimator carries practical consequences. Strategy 2a estimates each covariance element from pairwise complete observations and is straightforward to implement, but does not use the information in incomplete trajectories efficiently. In our simulations, this resulted in an empirical standard error about four times larger than for Strategy 2b in the realistic Scenario 3, and a small bias when dropout depended on observed values (Scenario 3b). Furthermore, the pairwise estimated covariance matrix is not guaranteed to be positive semi-definite, which occasionally led to correlation estimates outside [−1, 1]. As demonstrated in the ADNI case study, this can result in wider confidence intervals when dropout is substantial. With fewer than one third of subjects retained at the final timepoint, the pairwise complete-case estimate was essentially uninformative. Strategy 2b addresses this by fitting a linear mixed model to all available data, and yielded substantially narrower confidence intervals in the same application as well as unbiased estimates in all simulated scenarios. The drawback is that Strategy 2b depends on convergence of an iterative estimation procedure, which may fail under small sample sizes or near-singular covariance structures. In practice, we recommend Strategy 2b as the primary estimator when dropout is non-negligible, and Strategy 2a as a possible alternative when a sufficient number of complete cases is available. However, any other estimators of the variance-covariance matrix is possible to use in Strategy 2.

The results of the case study illustrate that the choice of strategy is not only a technical concern. Strategy 1 showed a near-zero correlation 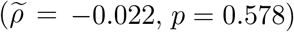, whereas Strategy 2b showed an arguably larger negative association 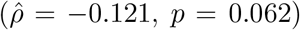, consistent with the expectation that faster brain aging is accompanied by greater grey matter loss. To be more explicit, both strategies might lead to different conclusions. We therefore recommend that researchers working with longitudinal neuroimaging data routinely inspect the missing data pattern and compute the weights as shown in Figure 4 to get an idea of how much each timepoint would be weighted when dropout is present. If the weight pattern seems not to be symmetric, we recommend using Strategy 2 instead over Strategy 1.

## Data Availability

Data used in preparation for this article were obtained from the Alzheimer's Disease Neuroimaging Initiative (ADNI) database (http://adni.loni.usc.edu).

## Acknowledgement

Ruben P. Dörfel was supported by a grant by the Fredrik och Ingrid Thurings Stiftelse (2025-207).

Data collection and sharing for this project was funded by the Alzheimer’s Disease Neuroimaging Initiative (ADNI) (National Institutes of Health Grant U01 AG024904) and DOD ADNI (Department of Defense award number W81XWH-12-2-0012). ADNI is funded by the National Institute on Aging, the National Institute of Biomedical Imaging and Bioengineering, and through generous contributions from the following: AbbVie, Alzheimer’s Association; Alzheimer’s Drug Discovery Foundation; Araclon Biotech; BioClinica, Inc.; Biogen; Bristol-Myers Squibb Company; CereSpir, Inc.; Cogstate; Eisai Inc.; Elan Pharmaceuticals, Inc.; Eli Lilly and Company; EuroImmun; F. Hoffmann-La Roche Ltd and its affiliated company Genentech, Inc.; Fujirebio; GE Healthcare; IXICO Ltd.; Janssen Alzheimer Immunotherapy Research & Development, LLC.; Johnson & Johnson Pharmaceutical Research & Development LLC.; Lumosity; Lundbeck; Merck & Co., Inc.; Meso Scale Diagnostics, LLC.; NeuroRx Research; Neurotrack Technologies; Novartis Pharmaceuticals Corporation; Pfizer Inc.; Piramal Imaging; Servier; Takeda Pharmaceutical Company; and Transition Therapeutics. The Canadian Institutes of Health Research is providing funds to support ADNI clinical sites in Canada. Private sector contributions are facilitated by the Foundation for the National Institutes of Health (www.fnih.org). The grantee organization is the Northern California Institute for Research and Education, and the study is coordinated by the Alzheimer’s Therapeutic Research Institute at the University of Southern California. ADNI data are disseminated by the Laboratory for Neuro Imaging at the University of Southern California.

## Conflict of Interest

Brice Ozenne states that part of his salary while working on this project was covered by a grant from Novo Nordisk A/S.

## A Extended Derivations

### A.1 Estimating *β*_*x*_ and *β*_*y*_ for each subject

For the simple linear regression *y*_*i,s*_ = *α*_*y,s*_ + *β*_*y,s*_*t*_*i*_ + *ε*_*i,s*_, we can estimate the slope 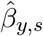 for each subject *s* using ordinary least squares:

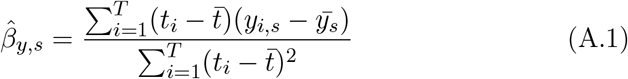

where the bar notation denotes the empirical average over time. Considering that the times *t* are fixed for all subjects and independent of *y*, we can rearrange (A.1) to

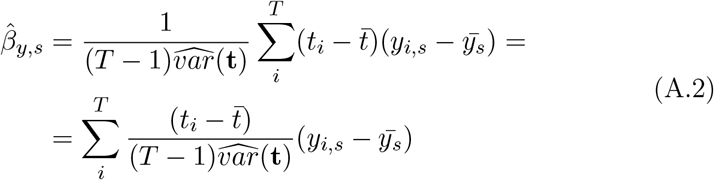

We can now introduce the weight *w*_*i*_

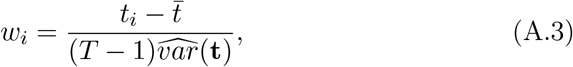

With

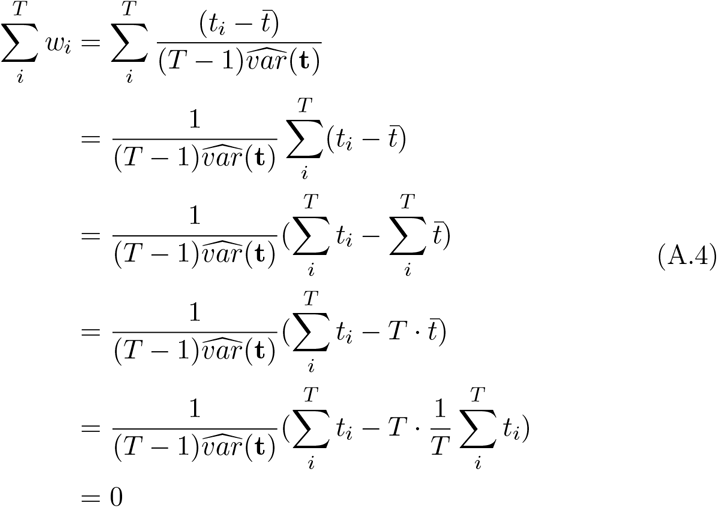

Please note that the weights *w*_*i*_ are independent of the measurement and are therefore applicable to both *x* and *y*. Substituting (A.3) into (A.2), while considering (A.4), we get

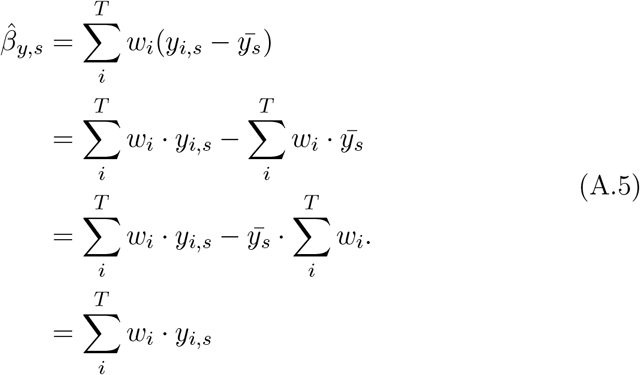

Similarly, we have 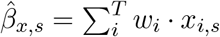 .

Now let **w** ∈ ℝ^*T* ×1^ be the vector containing all weights *w*_*i*_ and **Y, X** ∈ ℝ^**T**×**N**^ the matrices containing all values *y*_*i,s*_ and *x*_*i,s*_ respectively. We can therefore express 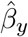 and 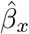both as 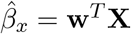 and 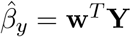.

### A.2 The expected value of *β*_*x*_ **and** *β*_*y*_

For reasons that will be obvious soon, we will start with the expected value of *y*_*i*_

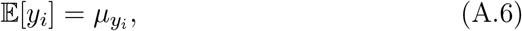

which can be estimated as

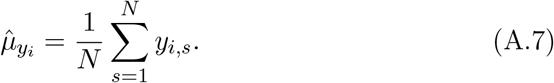

Similarly, the expected value of *β*_*y*_ is denoted:

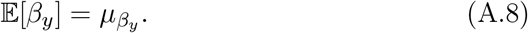

Now considering (A.5) and (A.7) we can estimate 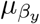as

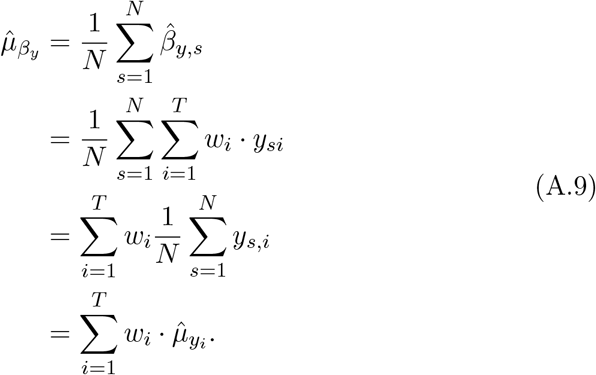

Similarly, we can get that 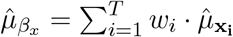

### A.3 The variance of *β*_*x*_ **and** *β*_*y*_

Following a similar approach to the previous subsection, we will start with the variance *var*(*y*_*i*_)

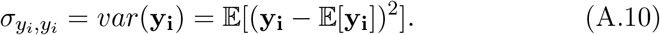

A suitable estimator would be

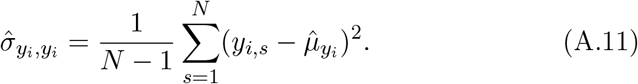

The variance 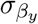, *β*_*y*_ is expected as

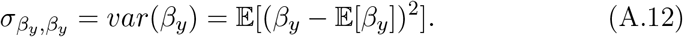

Utilizing (A.11), we would use the following estimator:

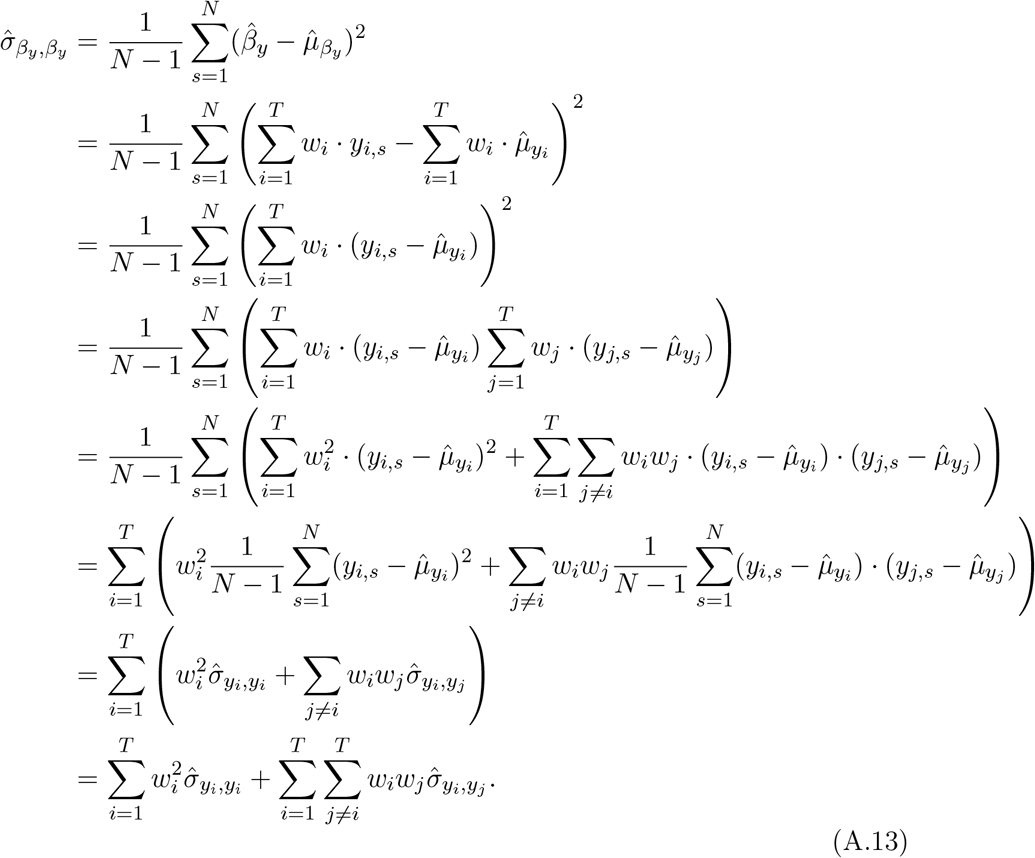

The first term sums the variances, and the second is the cross-terms of the empirical variance-covariance matrix 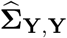. The keen reader might see that (A.13) is identical to 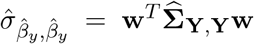 . By symmetry, we also have 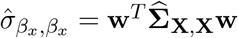.

### A.4 The covariance of *β*_*x*_ **and** *β*_*y*_

Since we never change a winning system, we start by deriving the expected value for the covariance 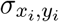 .

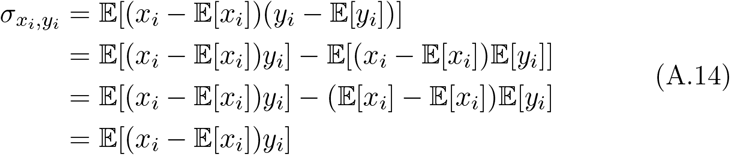

and provide a suitable estimator:

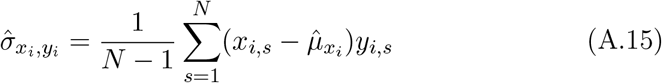

Moving on to the covariance 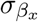, *β*_*y*_ we get:

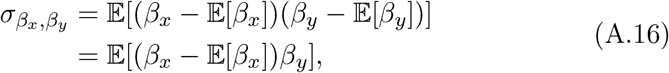

which can be estimated as:

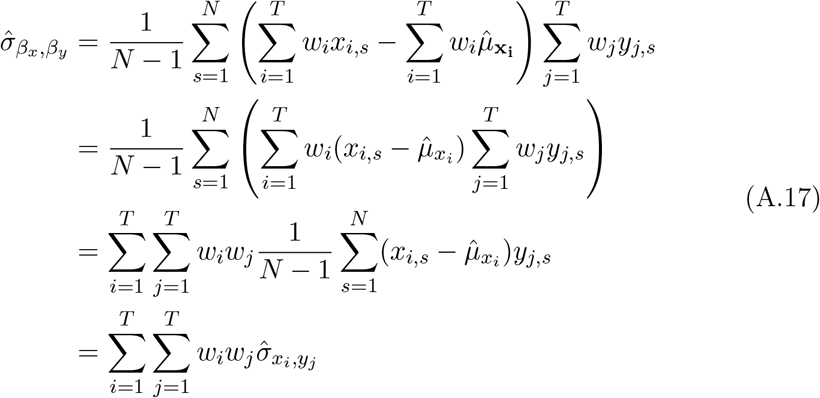

Again, the keen reader might see that eq. (A.17) is identical to 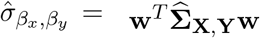.

In the case of correlating a baseline value *x*_1_ with longitudinal change *β*_*y*_, eq. (A.17) would simplify to

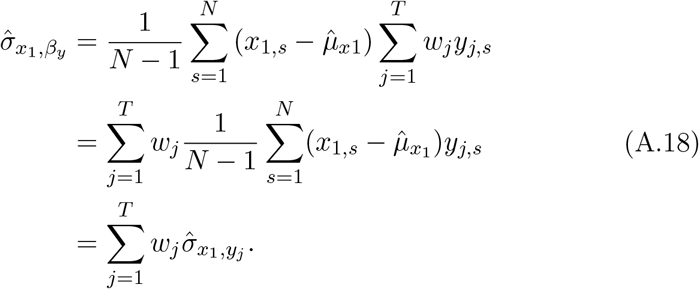

### A.5 Estimating 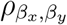

We now have estimated all the essential bits and pieces to estimate the correlation between the two slopes *β*_*x*_ and *β*_*y*_ (1). Essentially, we needed to estimate the elements of the variance-covariance matrix 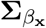, *β*_*y*_ . Considering (A.13) and (A.17) we get

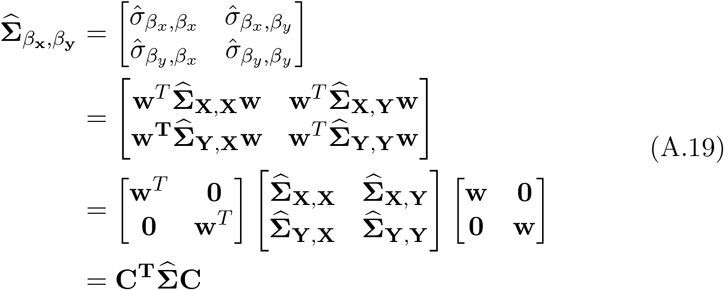

With **C** being the contrast matrix containing the weights. Therefore, it is possible to compute the correlation (1) directly from the contrast and the joined covariance matrix **Σ** of **X** and **Y**, without having to estimate the actual slopes for each subject. From this, we can see that both strategies for computing the correlation (1) are equivalent when all data is available. More specifically, estimating the respective slopes 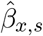 and 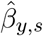 and their correlation is equivalent to calculating the joint covariance matrix **Σ** of **X** and **Y** and deducing the correlation from its elements:

### A.6 Equivalence between strategy 1 and strategy 2 with complete data

#### Equivalence between 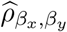 and 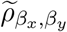 (no covariates)

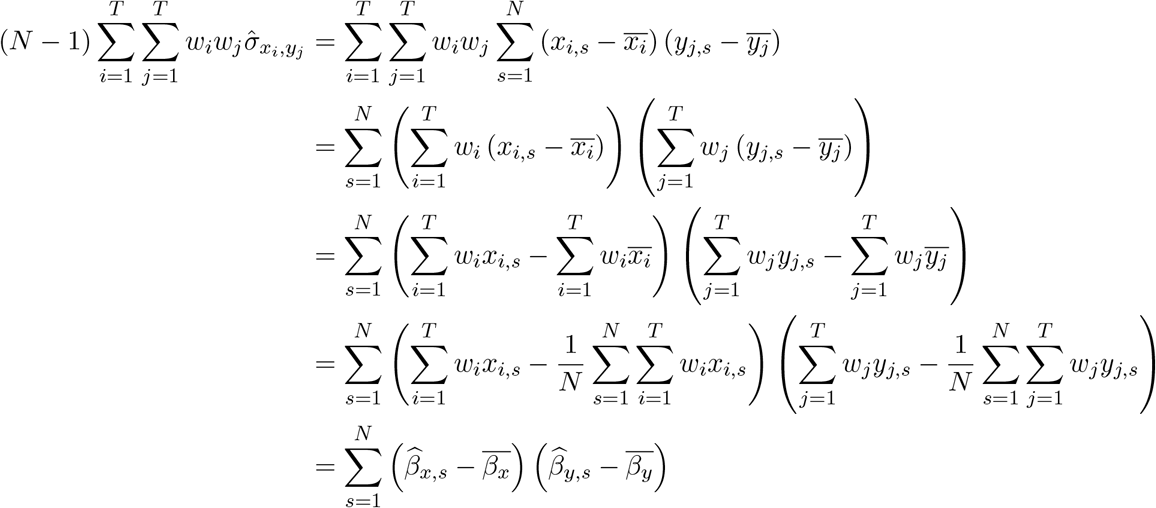

So the numerators of 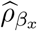, *β*_*y*_and 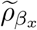, *β*_*y*_are equal. A similar argument can be used to show that the denominators are also equals and so are the two estimators.

#### Equivalence between 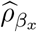, *β*_*y*_|*z* and 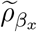, *β*_*y*_|z (covariates)

Denote by *I*_*N*_ the identity matrix and by *X*^(*Z*)^ = (*I* −*Z*(*Z*^*T*^ *Z*)^−1^*Z*^*T*^)*X* the projection of *X* in a space orthogonal to *Z*. The Frisch–Waugh–Lovell (FWL) theorem indicates that the estimated slope when adjusting on *Z* can be obtained by using *X*^(*Z*)^ instead of *X* when computing the slope using equation 3. Therefore:

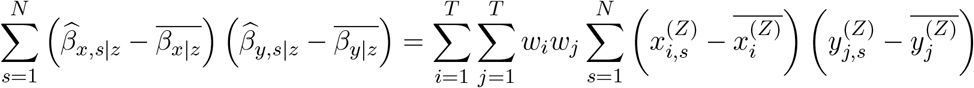

The conditional covariance is estimated as:

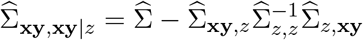

To simplify the derivation consider that all variables are centered and scaled, i.e., have mean 0 and variance 1. Using that the orthogonal projection *I*_*N*_ − **z z**^*T*^ **z z**^*T*^ is idempotent and symmetric, we have:

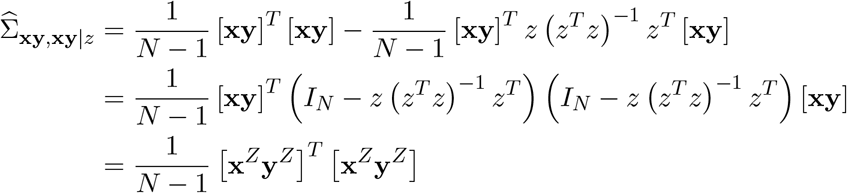

Therefore

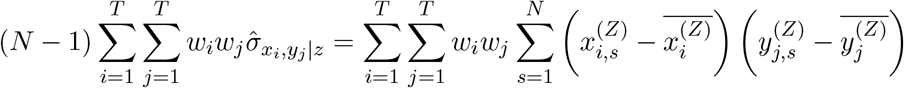

### A.7 Dropout

In presence of missing data and since we only sample at discrete time points, there is a finite amount (*K*) of possible patterns of missing data. Each missing data pattern correspond to a subset of the *T* measurement timepoints where we measure the two brain signals for *n*_*k*_ persons such that 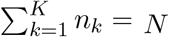 . We now show that, except in very special cases, the equivalence between the strategies 1 and 2 holds no more, even under independent dropout.

Example: consider the motivating example with *t* = 5 had there only been of (monotone) drop-out: subjects may leave the study before *τ* = 4. There are then *K* = 6 possible missing data pattern:

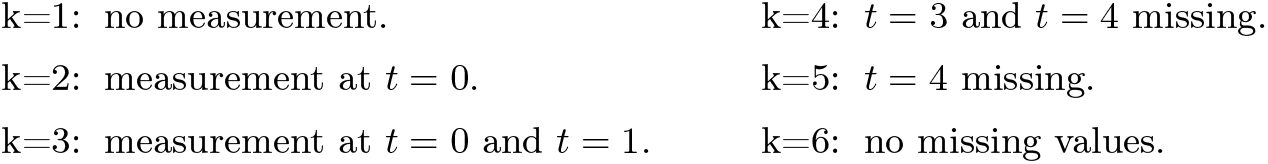

When estimating the subject-specific slope using strategy 1, we discard observations with fewer than two measurements and the weights are now dependent on the missing data pattern. Indeed, denote by *τ*_*k*_ the number of measurements under missing data pattern *k*, by 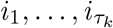 the index of the timepoints at which these measurements occurred, and by 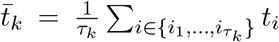 the average measurement time in missing data *k*. The estimated slope for the brain signal *y* is 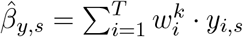 with:

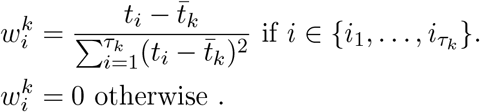

Example (continued): here *τ*_*k*_ = *k* − 1 and data with missing data pattern *k* = 1 and *k* = 2 are discarded since they have 0 or 1 measurement. For the other missing data patterns:

k=3: the estimated slope is 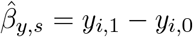 .

This follow from 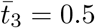 and 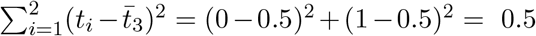 and that the only non-0 weights can be 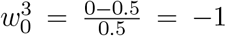 and 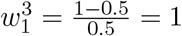.

k=4: the estimated slope is 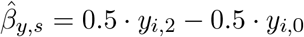 .

This follow from 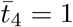 and 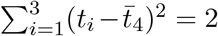 and that the only non-0 weights can be 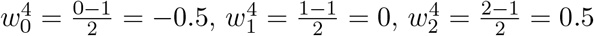 .

k=5: the estimated slope is 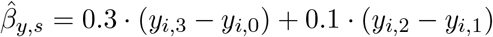 .

This follow from 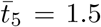 and 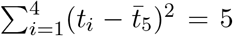 and that the only non-0 weights can be 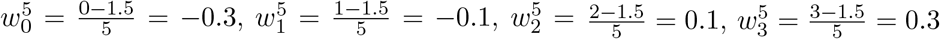 .

k=6: the estimated slope is 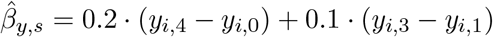 .

This follow from 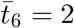 and 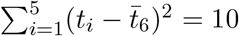 and that weights are 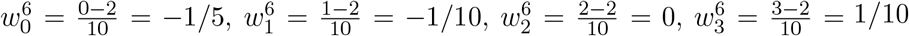, and 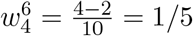 .

We can summarize the weights into a table:

| missing data pattern | $w_0$ | $w_1$ | $w_2$ | $w_3$ | $w_4$ |
| --- | --- | --- | --- | --- | --- |
| k=3 | -1 | 1 | 0 | 0 | 0 |
| k=4 | -0.5 | 0 | 0.5 | 0 | 0 |
| k=5 | -0.3 | -0.1 | 0.1 | 0.3 | 0 |
| k=6 | -0.2 | -0.1 | 0 | 0.1 | 0.2 |

A direct extension of the straightforward solution to accommodate missing values with strategy 1 is to then evaluate the Pearson correlation between the estimated slope. Following derivations similar to appendix A.3 and appendix A.4, we get:

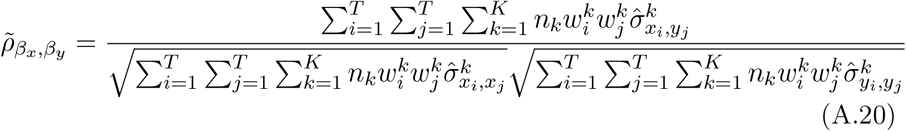

where 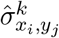 is the empirical covariance among individual with available *x* measurement at timepoint *t*_*i*_ and available *y* measurement at timepoint *t*_*j*_, i.e. same as estimator as 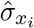,*y*_*j*_ but on a sub-population. In the case of independent dropout, i.e., subjects do not drop because they are ‘special’, but at random, and with a large enough sample in each subgroup, by consistency of the sample covariance estimator, we can assume that 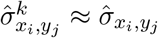, leading to:

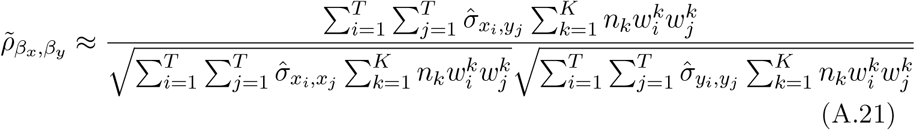

Example (continued): to further simplify consider the special case with equal number of subjects with each missing data pattern (*n*_*k*_ = *N/K*) and same missing data pattern for *x* and *y* (either both are measured or both are missing at a given timepoint). We display the value 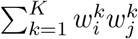 for each pair of timepoints (left table) and normalize it by the sum of the absolute value over all pairs _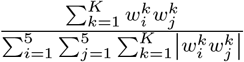_ (right table):

| $t_i \backslash t_j$ | 0 | 1 | 2 | 3 | 4 |
| --- | --- | --- | --- | --- | --- |
| 0 | 1.38 | -1.05 | -0.28 | -0.11 | -0.04 |
| 1 | -1.05 | 1.02 | 0.01 | 0.04 | 0.02 |
| 2 | -0.28 | 0.01 | 0.26 | 0.03 | 0.00 |
| 3 | -0.11 | 0.04 | 0.03 | 0.10 | 0.02 |
| 4 | -0.04 | 0.02 | 0.00 | 0.02 | 0.04 |
| 0 | 23.00 % | -17.50 % | -4.67 % | -1.83 % | -0.67 % |
| 1 | -17.50 % | 17.00 % | 0.17 % | 0.67 % | 0.33 % |
| 2 | -4.67 % | 0.17 % | 4.33 % | 0.50 % | 0.00 % |
| 3 | -1.83 % | 0.67 % | 0.50 % | 1.67 % | 0.33 % |
| 4 | -0.67 % | 0.33 % | 0.00 % | 0.33 % | 0.67 % |

which is to be compared to the case without missing values (i.e. missing data pattern 6): 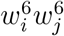 for each pair of timepoints (left table) and normalizing it by the sum of the absolute value over all pairs _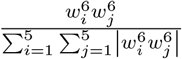_ (right table):

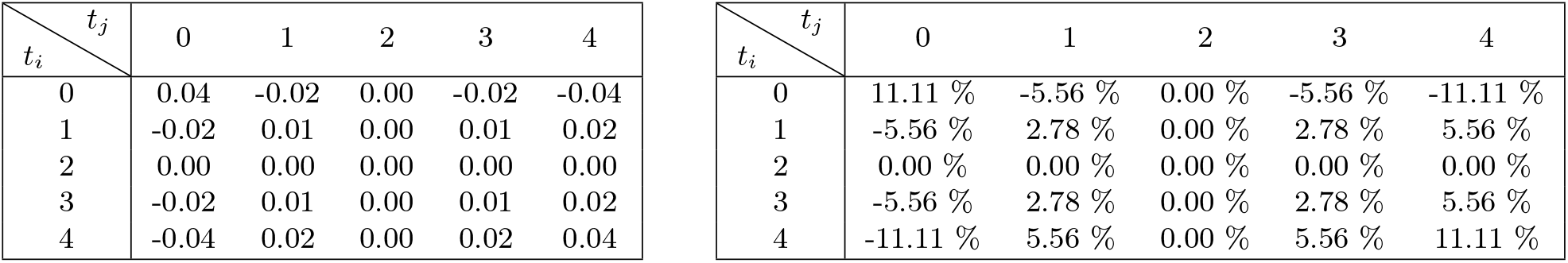

| $t_i \backslash t_j$ | 0 | 1 | 2 | 3 | 4 |
| --- | --- | --- | --- | --- | --- |
| 0 | 0.04 | -0.02 | 0.00 | -0.02 | -0.04 |
| 1 | -0.02 | 0.01 | 0.00 | 0.01 | 0.02 |
| 2 | 0.00 | 0.00 | 0.00 | 0.00 | 0.00 |
| 3 | -0.02 | 0.01 | 0.00 | 0.01 | 0.02 |
| 4 | -0.04 | 0.02 | 0.00 | 0.02 | 0.04 |
| 0 | 11.11 % | -5.56 % | 0.00 % | -5.56 % | -11.11 % |
| 1 | -5.56 % | 2.78 % | 0.00 % | 2.78 % | 5.56 % |
| 2 | 0.00 % | 0.00 % | 0.00 % | 0.00 % | 0.00 % |
| 3 | -5.56 % | 2.78 % | 0.00 % | 2.78 % | 5.56 % |
| 4 | -11.11 % | 5.56 % | 0.00 % | 5.56 % | 11.11 % |

This illustrates that the straightforward solution to accommodate missing values with strategy 1 puts more weight on pairwise covariance between early timepoints (1,2,3) and less weight on covariance involving later timepoints (4,5).

The above example illustrates that the 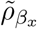, *β*_*y*_ correlation estimator is, except in special cases, a biased estimator of the correlation. Intuitively, it weights differently the pairs of timepoints leading to a different estimand whose definition is a function of the missing data pattern.

Denote by *I*_*T*_ the *T* × *T* identity matrix (1 on the diagonal and 0 otherwise) and by 1_*T*_ the *T* × *T* unit matrix (full of 1). When the variance-covariance for each brain signal and across brain signals has a compound symmetry structure:

- Σ_*X,X*_ = (1 − *ρ*_*x,x*_)*σ*_*x,x*_*I*_*T*_ + *ρ*_*x,x*_*σ*_*x,x*_1_*T*_ where *ρ*_*x,x*_ ∈] − 1, 1[
- Σ_*Y,Y*_ = (1 − *ρ*_*y,y*_)*σ*_*y,y*_*I*_*T*_ + *ρ*_*y,y*_*σ*_*y,y*_1_*T*_ where *ρ*_*y,y*_ ∈] − 1, 1[
- Σ_*X,Y*_ = (1 − *ρ*_*x,y*_)*σ*_*x,y*_*I*_*T*_ + *ρ*_*x,y*_*σ*_*x,y*_1_*T*_ where *ρ*_*x,y*_ ∈ [−1, 1].

the numerator of 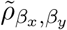 simplifies:

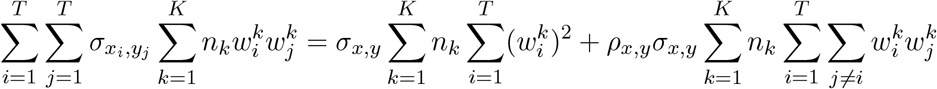

and similarly for the variance terms at the denominator. So when *ρ*_*x,y*_ = *ρ*_*x,x*_ = *ρ*_*y,y*_ = 0, the weight terms at the numerator and denominator will cancel out and 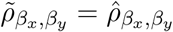 . This corresponds to scenario 1) in the simulations. When correlating a baseline measurement with a slope from another signal, it is enough that the covariance between the signal is constant over time to have that 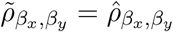 . Indeed in that case the estimated correlation will be 0 as 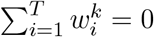 .

### A.8 Adjustment for Baseline Covariates

In many applications, we wish to estimate the correlation between changes in **X** and **Y** while adjusting for their baseline values, *x*_1_ and *y*_1_. This corresponds to estimating the partial correlation 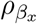, *β*_*y*_|*x*_1_,*y*_1_, which quantifies the association between slopes after accounting for baseline differences.

To achieve this, we extend the contrast matrix **C** to include baseline measurements. Let **e**_1_ ∈ ℝ^*T* ×1^ be a vector with 1 in the first position and 0 elsewhere, selecting the baseline timepoint. The adjusted contrast matrix is:

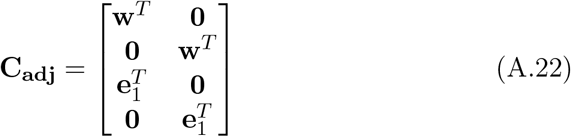

Applying this contrast to the joint covariance matrix yields:

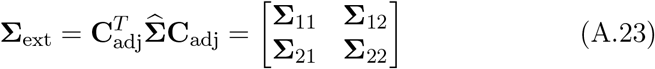

where **Σ**_11_ ∈ ℝ^2×2^ contains the covariances among *β*_*x*_ and *β*_*y*_, **Σ**_22_ ∈ ℝ^2×2^ contains the covariances among baseline values *x*_1_ and *y*_1_, and **Σ**_12_ contains the cross-covariances between slopes and baseline values.

Using the conditional covariance formula, the covariance matrix of the slopes adjusted for baseline is

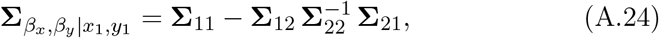

with the blocks

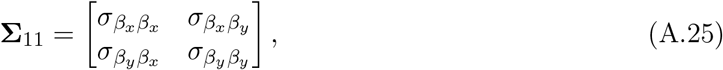

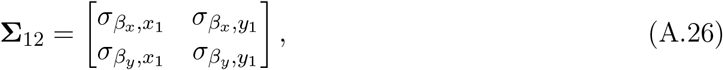

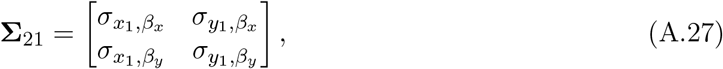

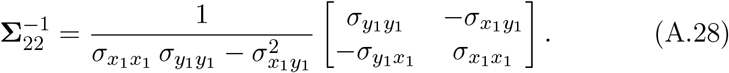

The individual entries can be written in terms of the weight vector **w** as

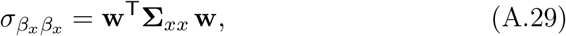

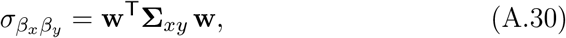

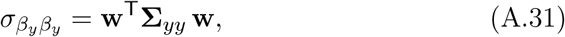

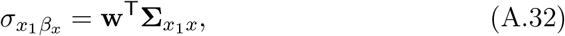

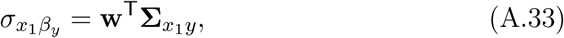

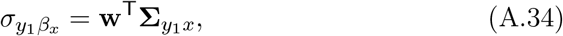

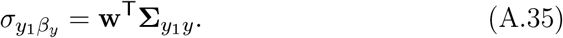

Substituting these scalar expressions into **Σ**_11_ gives

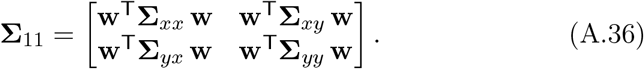

This can be written more compactly by introducing the block matrices

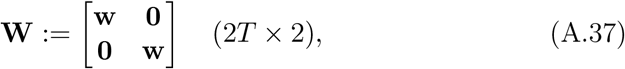

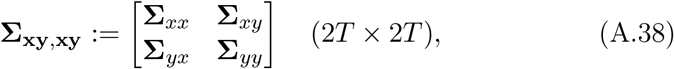

so that

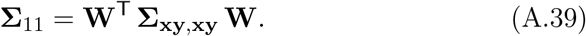

The same procedure applied to **Σ**_12_ yields

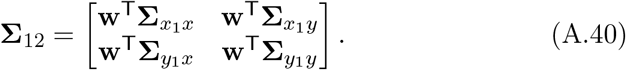

Defining the cross block

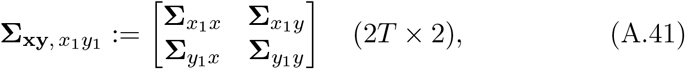

we obtain

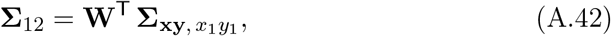

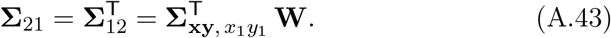

Substituting these compact forms into the conditional covariance formula gives

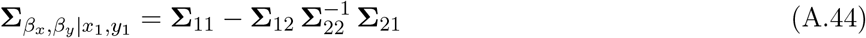

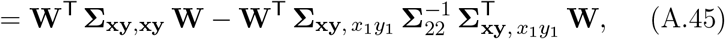

and factoring **W**^T^ on the left and **W** on the right yields

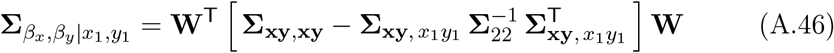

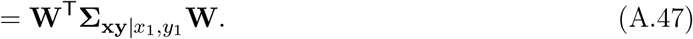

The partial correlation can then be computed as:

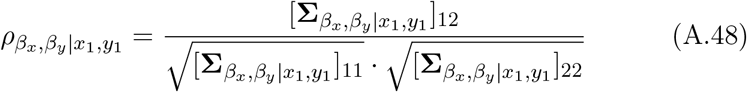

where 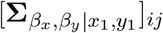 denotes the (*i, j*)-th element of the conditional covariance matrix. This approach naturally extends to adjusting for additional baseline covariates by expanding **C**_adj_ accordingly.

We can further show that when adjusting for baseline values, the marginal variance gets reduced by the variance explained by the baseline variables:

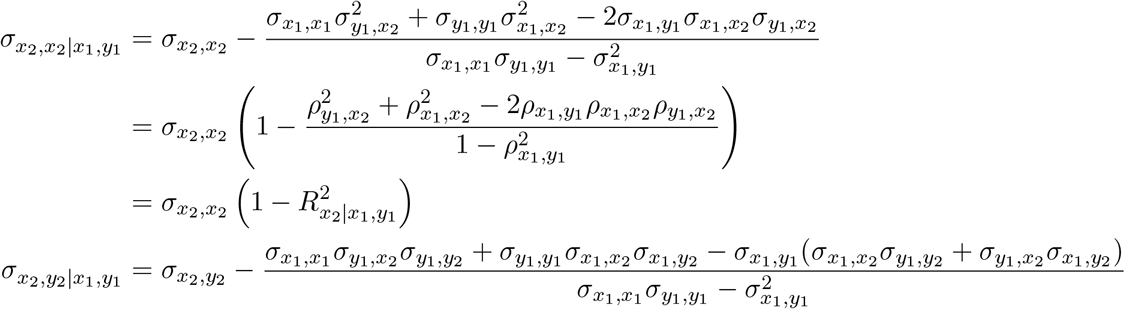

where 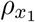,*y*_1_,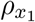,*x*_2_,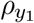,*x*_2_ denotes the pairwise correlation and 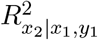 is the multiple correlation coefficient between *X*_2_ and the baseline signal values (*X*_1_, *Y*_1_) [9, Chapter 10]. We retrieve that 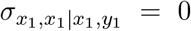 since 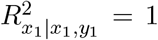 . To further simplify the conditional covariance we use that, after centering the outcome *X*_2_, the OLS estimator for the regression coefficients of *z* = (*X*_1_, *Y*_1_) can be estimated by 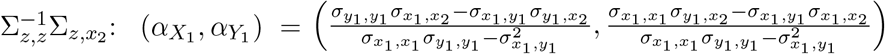 . . Therefore:

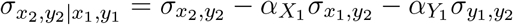

Since 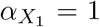 and 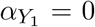 when *x*_2_ = *x*_1_, then 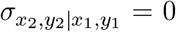 . The same result follow when *y*_2_ = *y*_1_ by symmetry between *x* and *y*.

**Figure B.1:**
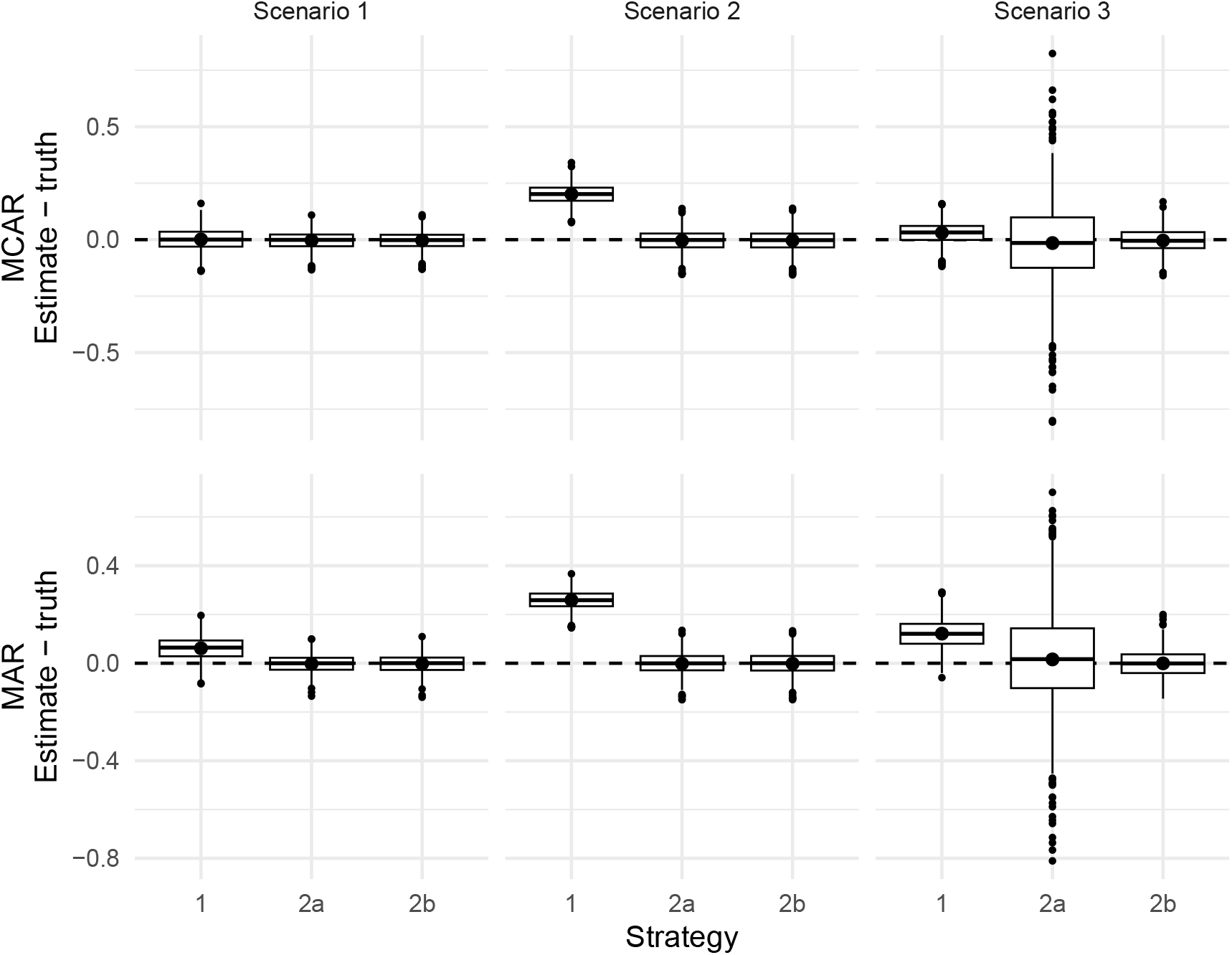
Error in estimated correlations between slopes (estimate -truth) across three covariance structures (Scenarios 1–3), with baseline adjustment. Top row: missing completely at random (MCAR); bottom row: missing at random (MAR) dropout. Strategy 1: slopes-first approach; Strategy 2a: joint covariance approach via pairwise complete case; Strategy 2b: joint covariance, approach via linear mixed model. The dashed line marks zero error (no bias). Boxplots show 1000 Monte Carlo replicates. The points show the mean error.

## B Extended Results

### B.1 Baseline adjustment

## Notes

### Summary of Updates

The author affiliations in the medRxiv interface have been updated. None of the files have changed.

